# Risk assessment of drug-induced Long QT Syndrome for some COVID-19 repurposed drugs

**DOI:** 10.1101/2020.04.21.20066761

**Authors:** Veronique Michaud, Pamela Dow, Sweilem B. Al Rihani, Malavika Deodhar, Meghan Arwood, Brian Cicali, Jacques Turgeon

## Abstract

The risk-benefit ratio associated with the use of repurposed drugs to treat 2019 SARS-CoV-2 related infectious disease (COVID-19) is complicated since benefits are awaited, not proven. A thorough literature search was conducted to source information on the pharmacological properties of 5 drugs and 1 combination (azithromycin, chloroquine, favipiravir, hydroxychloroquine, remdesivir, and lopinavir/ritonavir) repurposed to treat COVID-19. A risk assessment of drug-induced Long QT Syndrome (LQTS) associated with COVID-19 repurposed drugs was performed and compared to 23 well-known torsadogenic and 10 low torsadogenic risk compounds. Computer calculations were performed using pharmacokinetic and pharmacodynamic data, including affinity to block the rapid component of the delayed rectifier cardiac potassium current (I_Kr_) encoded by the human *ether-a-go-go* gene *(hERG)*, propensity to prolong cardiac repolarization (QT interval) and cause torsade de pointes (TdP). Seven different LQTS indices were calculated and compared. The U.S. Food and Drug Administration (FDA) Adverse Event Reporting System (FAERS) database was queried with specific key words relating to arrhythmogenic events. Estimators of LQTS risk levels indicated a very high or moderate risk for all COVID-19 repurposed drugs with the exception for azithromycin, although cases of TdP have been reported with this drug. There was excellent agreement among the various indices used to assess risk of drug-induced LQTS for the 6 repurposed medications and 23 torsadogenic compounds. Based on our results, monitoring of the QT interval shall be performed when some COVID-19 repurposed drugs are used, as such monitoring is possible for hospitalized patients or with the use of biodevices for outpatients.

## INTRODUCTION

As of August 13, 2020, there are 2,995 clinical trials registered for COVID-19, many assessing drugs to be repurposed for use against COVID-19; among those, hydroxychloroquine (N=216), azithromycin (N=94), lopinavir/ritonavir combination (N=67), chloroquine (N=74), dexamethasone (N=19), colchicine (N=18) and remdesivir (N=13).^1^ There have been a few promising preliminary results using some repurposed drugs in the therapeutic management of COVID-19, selected based upon their mechanism of action.^2^ Yet, the risk of doing so in COVID-19 patients has not been quantified, as a systematic approach should be used to assess risk benefit boundaries of these drugs. Furthermore, new safety profiles of repurposed drugs need to be established as they may have never been used or tested in such different patient populations. This is especially true for patients with complex drug regimen, polypharmacy, or treated with narrow therapeutic index drugs.

The American College of Cardiology (ACC) has issued a warning regarding the use of some of these drugs without evidence of whether their benefits outweigh their risks.^3^ Drugs like hydroxychloroquine and chloroquine prolong the QT interval, and may cause ventricular arrythmias and sudden cardiac arrest.^4-7^ We have recently conducted two risk assessment studies using simulations in specific populations of patients in whom these repurposed drugs could most likely be associated with unbalanced risk of cardiac toxicity (NCT04339634 and NCT04378881)^8,9^

This study looks at drugs proposed as potential COVID-19 therapies, assessing their risk of causing QT prolongation and their proarrhythmic properties that lead to a characteristic polymorphic ventricular tachycardia, described as torsade de pointes (TdP). As this study is based on literature review and computation from database information, patients’ specific characteristics, which are also important, could not be considered. Various risk assessments for drug-induced Long QT Syndrome (LQTS) will be presented and compared as predictive estimators for unbalanced risk benefit associated with repurposed drugs for COVID-19.

## METHODS

STEP 1: A literature search was conducted using the National Library of Medicine website (https://pubmed.ncbi.nlm.nih.gov/). A primary search used the key words “drug-induced QT prolongation index”. From this search, 38 publications were retrieved and considered for analysis. Six different indices were repeatedly referenced, including methodology and data on large numbers of drugs or elements to consider while evaluating the risk of drug-induced LQTS. The retained indices calculated and compared for drugs under study were based on reports from Redfern *et al*., 2003, Kramer *et al*., 2013, the Comprehensive *in vitro* Proarrhythmia Assay (CiPA), the CredibleMeds® website, Romero *et al*., 2018, and relative odds ratio calculated from the US Food and Drug Administration Adverse Event Reporting System (FAERS) database.^10-15^ We also included for comparison our developed and validated Long QT-JT index (information about calculation of this index and validation are described in details in Patent WO 2017/213825, United States of America, 2017).^16^

STEP 2: A second search was performed using the National Library of Medicine website (https://pubmed.ncbi.nlm.nih.gov/) with the following keywords (number of publications retrieved and analyzed are included in parentheses): “azithromycin and QT” (80), “azithromycin and torsade” (35), “azithromycin and arrhythmia” (97), “chloroquine and QT” (50), “chloroquine and torsade” (17), “chloroquine and arrhythmia” (178), “favipiravir and QT” (2), “favipiravir and torsade” (0), “favipiravir and arrhythmia” (1), “hydroxychloroquine and QT” (16), “hydroxychloroquine and torsade” (3), “hydroxychloroquine and arrhythmia” (58), “lopinavir/ritonavir and QT” (8), “lopinavir/ritonavir and torsade” (7), “lopinavir/ritonavir and arrhythmia” (19), “remdesivir and QT” (0), “remdesivir and torsade” (0), and “remdesivir and arrhythmia” (0).

STEP 3: A similar search was conducted by combining the key words “QT”, “torsade”, and “arrhythmia” with 19 “Known Risk”, 3 “Possible Risk” and 1 “Conditional Risk” torsadogenic drugs as reported by CredibleMeds® (https://www.crediblemeds.org/). These drugs were: “astemizole, chlorpromazine, cilostazol, cisapride, clarithromycin, clozapine, dasatinib, domperidone, donepezil, droperidol, escitalopram, halofantrine, haloperidol, lapatinib, methadone, ondansetron, pentamidine, pimozide, propofol, risperidone, terfenadine, thioridazine, and vandetanib”. The drugs were selected as relevant information was obtained for most indices under study. We also considered in our analysis 10 drugs with a low torsadogenic risk as described by Romero *et al*.^13^ These drugs are: Diltiazem, Duloxetine, Lamivudine, Loratadine, Mitoxantrone, Nifedipine, Raltegravir, Ribavirin, Sildenafil and Sitagliptin.

STEP 4: Pharmacological information [IC_50_ for hERG (KCNQ1; I_Kr_) block, IC_50_ for Nav1.5 (I_Na_) block, IC_50_ for Cav1.2 (I_Ca-L_) block, IC_50_ for Kv7.1 (KvLQT1; I_Ks_) block, peak plasma concentration (C_max_), maximum daily dose, plasma protein binding (%), inhibition of hERG trafficking, cardiac action potential duration (at 90% repolarization; APD_90_)] and clinical information (QT prolongation, torsade de pointe), retrieved from STEPs 2 and 3 were used to compute relevant QT indices retained from our analysis in STEP 1 for 6 COVID-19 repurposed medications, 23 known torsadogenic drugs and 10 low torsadogenic risk drugs. Data were either directly extracted or calculated using the reported equations from references identified in STEP 1. The calculated mean value was used as a parameter when more than one value has been reported in the literature. In the case of C_max_, the dose corrected value (or its mean) was extrapolated for the reported maximum daily dose. In the case of the Long QT-JT index, the following equations were used:

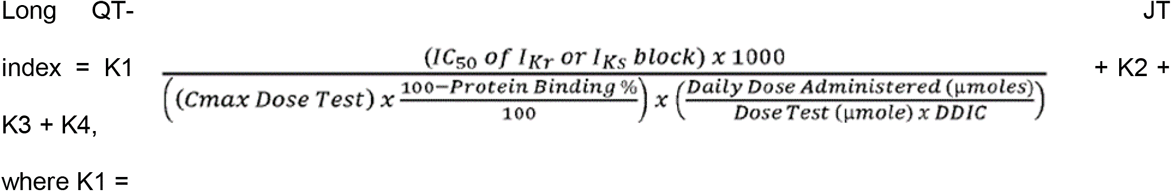

and DDIC is a drug-drug interaction coefficient calculated while considering the metabolic pathways of a drug being potentially inhibited by co-administered drugs. For drugs associated with block of I_Kr_ and I_Ks_, the lowest IC_50_ value was retained. As it pertains to DDIC, the partial metabolic clearance pathways have been characterized and estimated for each drug in the analysis. K1 is calculated under conditions of inhibition of the highest partial metabolic clearance. For example, if a drug has a CL_CYP2D6_ = 50%, CL_GYP2C19_ = 25%, CL_UGT1A9_ = 15% and a CL_renal_ = 10%, DDIC will consider a 50% possible decrease (CYP2D6) in the total clearance of this drug which will be reflected by a decreased K1 value by half; the lower the Long QT-JT index value, the higher is the risk.;

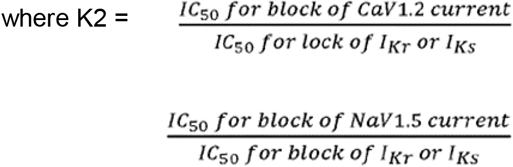

where K4 is given a value of −5 or 0 if the drug inhibits or does not inhibit hERG trafficking. K1, K2, K3 and/or K4 were set to 0 if no value is available for either IC_50_ for block of I_Kr_ or I_Ks_, IC_50_ for block of CaV1.2, IC_50_ for block of NaV1.5 or for hERG trafficking properties, respectively. STEP 5: SQL codes to perform queries on cleaned reports submitted to the FAERS database between 2004 and 2019Q2 were written for each of the 6 COVID-19 repurposed medications, 23 known torsadogenic drugs and 10 low torsadogenic drugs searching for adverse events reported for these drugs and a combination of specific search terms (see Table 3). An event was included in the final counts if the case report’s active ingredient, cleaned for text standardization and filtered by relevant route of administration for systemic exposure, and the reported preferred term were exact matches with the drug of interest and reference terms.

### Patient and Public Involvement

No patient involvement was included in this study. This study contains no observed drug-induced adverse effects in COVID-19 patients. Recommendations are based exclusively on searches of literature and databases.

## RESULTS

Results obtained for 7 indices of drug-induced LQTS, as well as the IC_50_ value for block of I_Kr_ (or I_Ks_), through the extraction of data from the literature or from computation using provided information for 6 COVID-19 medications are presented in Table 1. Using the same indices, the risk of these medications to induce LQTS can be compared to the risk measured for 23 well-established, torsadogenic drugs and 10 low torsadogenic drugs (Table 2). A color coded strategy is used to illustrate the correspondence between these various indices for well-know torsadogenic drugs, for drugs with low torsadogenic risk and for COVID-19 repurposed drugs. We also present the number of arrhythmogenic adverse drug events reported by the FAERS database for all drugs listed in Table 1 and 2 (Table 3). It is recognized that some bias certainly exists in the number of events reported by FAERS, as these events are not normalized for number of observed reports or prescriptions. However, FAERS data offer an appreciation of risk for drug-induced arrhythmogenic events in a “real-world” situation. In the following section, drug specific information will be provided to help appreciate and compare the risk of drug-induced LQTS for each of the 6 COVID-19 medications under consideration.

**Table 1.**
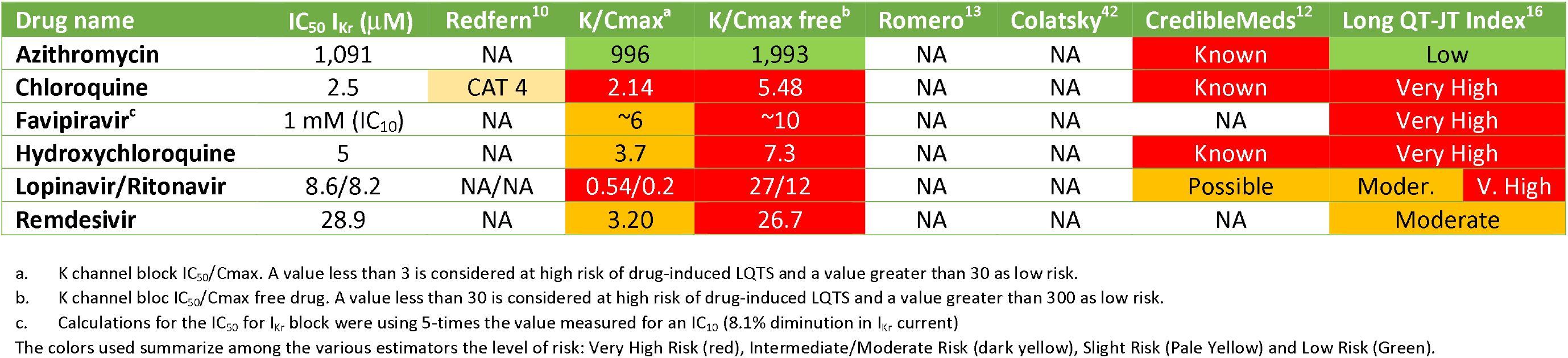
Relative indices of drug-induced LQTS risk associated with repurposed drugs used for COVID-19 treatment.

**Table 2.** Relative indices of risk associated with a series of drugs known to be associated with various risks for drug-induced LQTS.

| Drug name | IC<sub>50</sub> I<sub>Kr</sub> (μM) | Redfern<sup>10</sup> | K/Cmax<sup>a</sup> | K/Cmax free<sup>**b</sup> | Romero<sup>13</sup> | Colatsky<sup>42</sup> | CredibleMed<sup>12</sup> | Long QT-JT Index<sup>16</sup> |
| --- | --- | --- | --- | --- | --- | --- | --- | --- |
| Astemizole | 0.0012 | CAT 2 | 0.15 | 4.6 | Class 1 | Intermediate | Known | Very High |
| Chlorpromazine | 1.5 | CAT 3 | 1.4 | 39 | Class 1 | Intermediate | Known | Very High |
| Cilostazol | 13.8 | NA | 3.8 | 76 | Class 1 | NA | Known | Very High |
| Cisapride | 0.015 | CAT 2 | 0.12 | 5.8 | Class 1 | Intermediate | Known | Very High |
| Clozapine | 2.3 | NA | 1.0 | 33 | Class 2 | Intermediate | Possible | Very High |
| Domperidone | 0.057 | CAT 4 | 0.17 | 2.2 | Class 1 | Intermediate | Known | Very High |
| Droperidol | 0.028 | CAT 2 | 0.16 | 1.8 | Class 1 | Intermediate | Known | Very High |
| Halofantrine | 0.38 | CAT 3 | 0.03 | 2.2 | Class 1 | NA | Known | Very High |
| Haloperidol | 0.04 | CAT 3 | 0.75 | 10.0 | Class 1 | NA | Known | Very High |
| Methadone | 3.5 | NA | 0.86 | 6.9 | Class 1 | NA | Known | Very High |
| Ondansetron | 0.81 | NA | 1.2 | 4.9 | Class 1 | Intermediate | Known | Very High |
| Pentamidine | 1.28 | CAT 3 | 1.7 | 5.3 | NA | NA | Known | Very High |
| Pimozide | 0.015 | CAT 3 | 0.28 | 30 | NA | Intermediate | Known | Very High |
| Propofol | 30 (IKs) | NA | 1.7 | 57 | NA | NA | Known | Very High |
| Risperidone | 0.261 | CAT 5 | 2.1 | 21 | Class 2 | Intermediate | Conditional | Very High |
| Terfenadine | 0.016 | CAT 2 | 1.7 | 55 | Class 1 | Intermediate | Known | Very High |
| Thioridazine | 0.500 | CAT 3 | 0.28 | 0.51 | Class 1 | NA | Known | Very High |
| Vandetanib | 0.400 | NA | 0.09 | 0.94 | NA | High | Known | Very High |
| Clarithromycin | 39.3 | CAT 4 | 12 | 41 | Class 1 | Intermediate | Known | Moderate |
| Donepezil | 0.7 | NA | 11 | 233 | Class 1 | NA | Known | Moderate |
| Escitalopram | 2.6 | NA | 28 | 64 | NA | NA | Known | Moderate |
| Lapatinib | 1.1 | NA | 0.26 | 53 | Class 2 | NA | Possible | Moderate |
| Dasatinib | 24.5 | NA | 24 | 598 | Class 2 | NA | Possible | Low |
| Diltiazem | 13.2 | CAT 5 | 24 | 96 | Class 4 | Low | NA | Low |
| Duloxetine | 3.8 | NA | 23 | 238 | Class 4 | NA | NA | Low |
| Lamivudine | 2054 | NA | 67 | 105 | Class 4 | NA | NA | Low |
| Loratadine | 6.1 | CAT 5 | 260 | 12976 | Class 4 | Low | NA | Low |
| Mitoxantrone | 539.4 | NA | 521 | 2369 | Class 4 | NA | NA | Low |
| Nifedipine | 44.0 | CAT 4 | 227 | 5686 | Class 4 | Low | NA | Low |
| Raltegravir | 782.8 | NA | 19 | 112 | Class 4 | NA | NA | Low |
| **Ribavirin** | 967 | NA | 86 | 86 | Class4 | NA | NA | Low |
| **Sildenafil** | 33.3 | NA | 35.5 | 878 | Class4 | NA | NA | Low |
| **Sitagliptine** | 174.7 | NA | 245 | 396 | Class 4 | NA | NA | Low |

a. K channel block $IC_{50}/C_{max}$ . A value less than 3 is considered at high risk of drug-induced LQTS and a value greater than 30 as low risk.
b. K channel block $IC_{50}/C_{max}$ free drug. A value less than 30 is considered at high risk of drug-induced LQTS and a value greater than 300 as low risk.
The colors used summarize among the various estimators the level of risk: Very High Risk (red), Intermediate/Moderate Risk (dark yellow), Slight Risk (Pale Yellow) and Low Risk (Green). Drugs are ordered alphabetically based on their Long QT-JT index classification from very high to low risk.

**Table 3.** Number of arrhythmogenic adverse drug events reported by the FDA Adverse Event Reporting System for repurposed drugs used for COVID-19 treatment and drugs known to be associated with various risk for drug-induced LQTS.

| DRUG NAME<sup>a</sup> | All terms<sup>b</sup> | VT/VF/TdP/LQTS<sup>c</sup> | VT/VF/VA/VFL/VT<sup>d</sup> | TdP/LQTS<sup>e</sup> | CA/C-RA<sup>f</sup> |
| --- | --- | --- | --- | --- | --- |
| **COVID-19 DRUGS** |  |  |  |  |  |
| AZITHROMYCIN | 790 | 168 | 115 | 53 | 171 |
| CHLOROQUINE | 129 | 40 | 28 | 12 | 36 |
| HYDROXYCHLOROQUINE | 240 | 73 | 38 | 35 | 48 |
| LOPINAVIR/RITONAVIR | 501 | 42 | 30 | 12 | 90 |
| RITONAVIR | 206 | 35 | 28 | 7 | 81 |
| **HIGH RISK DRUGS** |  |  |  |  |  |
| CHLORPROMAZINE | 48 | 0 | 3 | 0 | 9 |
| CILOSTAZOL | 298 | 60 | 79 | 20 | 52 |
| CISAPRIDE | 1026 | 162 | 173 | 77 | 38 |
| CLOZAPINE | 2530 | 29 | 55 | 6 | 686 |
| DOMPERIDONE | 21 | 5 | 7 | 1 | 0 |
| DROPERIDOL | 26 | 7 | 11 | 2 | 3 |
| HALOFANTRINE | 4 | 1 | 1 | 1 | 1 |
| HALOPERIDOL | 723 | 144 | 81 | 100 | 176 |
| METHADONE | 1537 | 220 | 114 | 174 | 661 |
| ONDANSETRON | 854 | 191 | 184 | 125 | 192 |
| PENTAMIDINE | 2 | 0 | 0 | 0 | 1 |
| PIMOZIDE | 33 | 7 | 6 | 4 | 7 |
| PROPOFOL | 675 | 133 | 198 | 31 | 348 |
| RISPERIDONE | 1608 | 131 | 108 | 67 | 324 |
| THIORIDAZINE | 56 | 6 | 9 | 1 | 10 |
| VANDETANIB | 93 | 1 | 1 | 1 | 2 |
| **MODERATE RISK DRUGS** |  |  |  |  |  |
| CLARITHROMYCIN | 769 | 182 | 113 | 118 | 127 |
| DONEPEZIL | 1108 | 157 | 103 | 121 | 172 |
| ESCITALOPRAM | 838 | 56 | 59 | 28 | 128 |
| LAPATINIB | 251 | 7 | 8 | 3 | 47 |
| **LOW RISK DRUGS** |  |  |  |  |  |
| DASATINIB | 180 | 7 | 9 | 2 | 57 |
| DILTIAZEM | 725 | 87 | 55 | 32 | 385 |
| DULOXETINE | 1890 | 92 | 82 | 10 | 156 |
| LORATADINE | 338 | 74 | 40 | 34 | 32 |
| NIFEDIPINE | 216 | 36 | 33 | 3 | 93 |
| RALTEGRAVIR | 81 | 25 | 23 | 2 | 20 |
| RIBAVIRIN | 581 | 27 | 23 | 4 | 72 |
| SILDENAFIL | 856 | 131 | 127 | 4 | 220 |

- a. No data are available in the FAERS database for favipiravir, remdesivir, astemizole, terfenadine, lamivudine, mitotraxone and sitagliptine. - b. "All terms" refers to the following terms included in the query: Electrocardiogram QT prolonged, Long QT syndrome, Long QT syndrome congenital, Torsade de pointes, Ventricular tachycardia, Cardiac arrest, Cardiac death, Cardiac fibrillation, Cardio-respiratory arrest, Electrocardiogram repolarization abnormality, Electrocardiogram U wave inversion, Electrocardiogram U wave present, Electrocardiogram U-wave abnormality, Loss of consciousness, Multiple organ dysfunction syndrome, Sudden cardiac death, Sudden death, Syncope, Ventricular arrhythmia, Ventricular fibrillation, Ventricular flutter, Ventricular tachyarrhythmia. - c. Terms included in the query comprised: Ventricular Tachycardia (VT), Ventricular fibrillation (VF), Torsade de pointes (TdP), Long QT Syndrome (LQTS), Ventricular arrhythmia, ventricular flutter, Cardiac fibrillation. - d. Terms included in the query comprised: Ventricular Tachycardia (VT), Ventricular fibrillation (VF), Ventricular arrhythmia (VA), Ventricular flutter (VFL), Cardiac fibrillation (CF). - e. Terms included in the query comprised: Long QT Syndrome (LQTS), Torsade de pointes (TdP) - f. Terms included in the query comprised: Cardiac arrest (CA), Cardio-respiratory arrest (C-RA)

### Azithromycin

Azithromycin use has been proposed in conjunction with chloroquine or hydroxychloroquine to prevent or treat concomitant bacterial infections in patients with COVID-19.^17^ Peak plasma concentrations in the range of 400 ng/mL (550 nM) are observed following a single 500 mg oral dose.^18, 19^

*QT prolongation and TdP*. Prolongation of cardiac repolarization and QT interval, imparting a risk of developing cardiac arrhythmia and TdP, has been reported in patients treated with macrolides.^20^ All estimators of azithromycin-related risk of drug-induced TdP (Table 1) suggest a weak proarrhythmic effect of the drug (IC_50_ for block of hERG between 0.856 mM and 1.091 mM), although cases of TdP have been described.^21^ Furthermore, cases of TdP have been spontaneously reported during post-marketing surveillance (FAERS) in patients receiving azithromycin (Table 3). This explains why CredibleMeds® lists azithromycin as a “Known Risk” drug.

### Chloroquine

Chloroquine is metabolized by CYP2C8 and CYP3A4 through dealkylation to monedethylchloroquine and bisdesethylchloroquine.^22^ Peak plasma concentrations of chloroquine in the range of 1.0-3.0 μM have been measured following the oral administration of a 600 mg oral dose to healthy subjects or patients.^23, 24^

*QT prolongation and TdP*. Chloroquine has been shown to block hERG channels with an IC_50_ of 2.5 μM.^4^ Redfern *et al*. described chloroquine as a Category 4 drug, *i.e*., drugs for which there have been isolated reports of TdP in humans.^10^ Indeed, case reports of chloroquine-induced QT prolongation, cardiac electrophysiology disturbance and TdP have been reported. Based on CredibleMeds® classification, chloroquine is a drug of “Known Risk” for TdP.^25^ The Long QT-JT index calculated for chloroquine under conditions of CYP2C8 inhibition is Very High (Table 1), a risk-estimate value similar to that calculated for drugs such as astemizole, haloperidol, pimozide, and terfenadine (Table 2).^16^

### Favipiravir

Favipiravir is a new antiviral drug with a broad activity toward RNA viruses, such as the influenza virus, including the avian influenza (H5N1), rhinovirus, and respiratory syncytial virus.^26, 27^ The usual adult dosage is 1,600 mg of favipiravir administered orally twice daily on day one, followed by 600 mg orally twice daily from day two through day five. Favipiravir C_max_ in plasma is about 500 μM (78.9 μg/mL; 1,600 mg dose). It could also inhibit CYP2C8 with an IC_50_ of 477 μM (74.9 μg/ml).^28^

*QT prolongation and TdP*. Studies looking at the effects of favipiravir on hERG block have shown that a mild suppression of the hERG current was observed at the concentration of 1 mM (157 μg/mL, 2.0 times the observed human C_max_ at a dose of 1,600 mg). In a telemetry study conducted in dogs, no effects on blood pressure (systolic, diastolic, mean), heart rate, or ECG parameters (PR interval, QRS, QT and QTc) were observed even at the oral dose of 150 mg/kg [C_max_ (mean), 268 μg/mL; 3.4 times the human C_max_)] until 20 hours have passed after the treatment.^28^ Calculation of K/C_max_, K/C_max_ free and Long QT-JT Index using an estimated IC_50_ of 5 mM (5-times the concentration associated with 8.1% block of I_Kr_) yielded parameters in the high- to very high-risk categories for drug-induced LQTS. As no reports of LQTS have been published yet, close monitoring of QTc remains advisable.

### Hydroxychloroquine

Major metabolic pathway is through N-dealkylation leading to the formation of desethylhydroxychloroquine; desethylchloroquine and bisdesethylchloroquine are minor metabolites. The N-desalkylation pathway seems to be mediated by CYP2C8, CYP3A4, and CYP2D6 (high affinity, but low capacity). Following a single 200 mg oral dose to healthy males, the plasma C_max_ was 50.3 ng/mL (150 nM), reached in 3.74 hours with an elimination half-life of 123.5 days. Administration of hydroxychloroquine 200 mg three times a day led to mean serum C_max_ of 460 ng/mL (1,3 μM).^17^

*QT prolongation and TdP*. Use of hydroxychloroquine for the treatment of lupus erythematosus has been associated with occasional reports of QT interval prolongation and TdP.^7^ In their recent study using hydroxychloroquine in combination with azithromycin, Chorin *et al*., observed QTc increases of 40 msec or more in 30% of their patients; in 11% of their 84 patients, QTc increased to more than 500msec.^6^ CredibleMeds® has classified hydroxychloroquine as a “Known Risk” drug for TdP. K/C_max_, K/C_max_ free and Long QT-JT index also identify hydroxychloroquine as a high-risk drug for drug-induced LQTS.

### Lopinavir/ritonavir

When administered alone, lopinavir is a low affinity CYP3A4 substrate with a low oral bioavailability. As a high affinity substrate of CYP3A4, ritonavir is co-administered to inhibit the first pass metabolism of lopinavir and increases its plasma concentrations by about 10-fold. Under such conditions, lopinavir reaches about 10 μg/ml (15.9 μM) after an 800 mg dose. For its part, ritonavir has a bioavailability of 85%, and C_max_ of about 11 μg/mL (15.5 μM) are observed after administration of a 600 mg oral dose.^29^ Importantly, because of strong CYP3A4 inhibition, major drug-drug interactions are observed with other CYP3A4 substrates.^30-32^

*QT prolongation and TdP*. Lopinavir and ritonavir are associated with significant blockade of the hERG (I_Kr_) channel^33^ (Table 1) and cases of TdP have been registered in the FAERS database (Table 3). The monograph of the product also states that cases of QT interval prolongation and TdP have been reported, although causality of lopinavir/ritonavir combination could not be established. CredibleMeds® lists the drug under the category “Possible Risk”, while other indices list this combination under High Risk (Table 1).

### Remdesivir

Remdesivir is an investigational antiviral drug. Originally tested in Ebola patients, it is not currently FDA-approved to treat or prevent any diseases, including COVID-19.^34, 35^ However, the FDA has approved an Emergency Use Authorization (EUA) to allow treatment of suspected or laboratory confirmed COVID-19 in adults and pediatric patients hospitalized with severe disease. In addition, remdesivir was recently approved by the Japanese Ministry of Health, Labor and Welfare (MHLW) to treat COVID-19 infection. Remdesivir also received a provisional acceptance by the Therapeutic Goods Administration (TGA) of Australia to be used in adults and adolescents hospitalized with severe COVID-19 symptoms.

Finally, the European Medicines Agency (EMA) recently recommended granting a conditional marketing authorization to remdesivir for the treatment of COVID-19 in adults and adolescents from 12 years of age with pneumonia who require supplemental oxygen. *In vitro*, remdesivir undergoes metabolism by CYP2C8, CYP2D6, and CYP3A4. It is not suitable for oral delivery because of its poor hepatic stability.^36^ Peak plasma concentrations reached 9.0 μM following administration of a 200 mg intravenous infusion over 30 min.^36^

*QT prolongation and TdP*. Very little is known at this stage about the risk of drug-induced LQTS by remdesivir. An IC_50_ of 28.9 mM has been estimated *in vitro* for block of hERG (I_Kr_) (Table 1). Calculated values for K/C_max_, K/C_max_ free, and Long QT-JT index indicate that remdesivir use could carry a significant risk, especially if its intravenous administration leads to high peak plasma levels. Therefore, close monitoring on the QT interval appears warranted.

## DISCUSSION

Based on our literature search and use of information computed from databases, several currently proposed drugs to be used off-label and repurposed for the treatment of COVID-19 are associated with a significant risk of drug-induced LQTS. Therefore, mandatory monitoring of the QT interval should be performed. Such monitoring could be easily performed for hospitalized patients, but would require the use of biodevices for outpatients initiated on these drugs.

Several approaches have been proposed to assess risk of drug-induced LQTS. Among the most important were the S7B and E14 International Conference on Harmonisation Guidances for non-clinical and clinical evaluation of new non-antiarrhythmic human pharmaceuticals.^37, 38^ After the introduction of these guidelines in 2005, no drugs were removed from the market due to TdP risk. However, concerns raised about the optimal fail-fast fail-safe paradigm in recent drug development programs led to unnecessary removal of promising therapeutic molecules.^39^ There were also concerns regarding the financial burden associated with these types of studies; therefore, alternative approaches were evaluated.^40^

In 2003, Redfern *et al*. published an exhaustive review on a broad range of drugs looking at hERG (I_Kr_) activity, cardiac action potential duration (at 90% repolarization; APD_90_), and QT prolongation in dogs.^10^ They compared these properties against QT prolonging effects of drugs and reports of TdP in humans. The investigators considered the free plasma concentrations attained during clinical use and classified drugs into five categories. Category 1 includes repolarization-prolonging antiarrhythmics; Category 2 includes drugs that have been withdrawn or suspended from the market in at least one major regulatory territory due to an unacceptable risk of TdP for the condition being treated; Category 3 includes drugs that have a measurable incidence of TdP in humans, and those for which numerous case reports exist in the published literature; Category 4 includes drugs for which there have been isolated reports of TdP in humans; and Category 5 includes drugs for which there have been no published reports of TdP in humans.

In 2013, Kramer *et al*. assessed concomitant block of multiple ion channels (Multiple Ion Channel Effects, MICE) by measuring the concentration-responses of hERG (I_Kr_), Nav1.5 (I_Na_) and Cav1.2 (I_Ca-L_) currents for 32 torsadogenic and 23 non-torsadogenic drugs from multiple pharmacological classes.^11^ The best logistic regression models using the MICE assay only required a comparison of the blocking potencies between hERG and Cav1.2. Unfortunately, drug-specific indices or values associated with their drug comparison model were not provided. Other much simpler indices, such as K/C_max_, *i.e*., IC_50_, for block of I_Kr_/peak plasma concentration (C_max_ at maximum dose) or K/ C_max_ free, *i.e*. IC_50_ for block of I_Kr_/C_max_ unbound concentration (considering plasma protein binding) have also been proposed.^10^ Values ≤3 and ≤30 for these respective factors are generally considered indicative of high risk of drug-induced LQTS.

More recently, the Comprehensive *in vitro* Proarrhythmia Assay (CiPA) initiative was established by a partnership of several organizations. Their main objective was to develop a new paradigm for assessing proarrhythmic risk, building on the emergence of new technologies and an expanded understanding of torsadogenic mechanisms beyond I_Kr_ block.^41^ Their strategy involves the use of three pillars to evaluate drug effects: 1) human ventricular ionic channel currents in heterologous expression systems, 2) *in silico* integration of cellular electrophysiologic effects based on ionic current effects, and 3) fully integrated biological systems (stem-cell-derived cardiac myocytes and the human ECG). In their most recent publication, they reported on a list of 28 drugs under three risk categories.^42^ The High TdP Risk category includes mostly Class III or Class 1A antiarrhythmics; the Intermediate Risk category groups drugs that are generally accepted to be torsadogenic; and the Low Risk category included drugs known not to be associated with TdP.

Other groups have concentrated their efforts on creating *in silico* tools for the early detection of drug-induced proarrhythmic risks.^13, 43^ For instance, Romero *et al*. looked at the effects of drugs on action potential duration of isolated endocardial, myocardial, and epicardial cells as well as the QT prolongation in virtual tissues using multiple channel-drug interactions and state-of-the-art human ventricular action potential models.^44^ Based on 206,766 cellular and 7,072 tissue simulations assessing block of I_Kr_, I_Ks_ and I_Ca-L_, they studied 84 compounds and classified 40 of them as torsadogenic. They proposed the use of a new index, Tx, for differentiating torsadogenic compounds. Tx was defined as the ratio of the drug concentrations producing 10% prolongation (similar to an IC_10_ rather than IC_50_) of the cellular endocardial, midmyocardial, and epicardial APDs, and the QT interval, over the maximum effective free therapeutic plasma concentration.

For many years, a group led by Dr. Raymond Woosley has accumulated information on drugs associated with LQTS.^25^ CredibleMeds® reviews available evidence for these drugs and places them into one of three designated categories: “Known Risk” of TdP, “Possible Risk” of TdP, and “Conditional Risk” of TdP. They have also created a list of drugs to avoid in patients with genetically inherited LQTS. The merit of CredibleMeds® is the classification of drugs based on clinically observed and documented cases of TdP of QT prolongation.

Another approach to assess risk of drug-induced LQTS is to use large collections of individual and manufacturer-reported adverse drug events. One of the most popular resources for these types of reports is the FDA Adverse Event Reporting System (FAERS), a data repository developed to support the FDA’s post-marketing safety surveillance program. This program is charged with monitoring the occurrence and safety signals of approximately 104 adverse events for drugs and therapeutic biologic products, as standardized by the Medical Dictionary of Regulatory Affairs. The FAERS repository contains information on adverse drug events and medication error reports submitted to the FDA, and makes these reports publicly available on a quarterly basis. Although it presents some limitations, the appropriate adjustment of this data allows the capture of signals suggesting adverse drug events associated with a particular drug. On an individual and/or population basis, FAERS data can be used to identify patients who are at highest risk of adverse drug events using statistical approaches such as relative odds.

All of the sus-mentioned approaches are complementary while each having their own weaknesses and strengths. Recognizing the complexity of determining the risk of drug-induced LQTS noted by others ^45^, our team has developed the Long QT-JT index, which uses algorithms that consider IC_50_ for block of relevant ion channels (I_Kr_, I_Ks_, I_Na_, I_Ca-L_), inhibition of hERG trafficking, unbound C_max_ at maximum dose, and most importantly the inhibition of the major metabolic pathway involved in the disposition or torsadogenic drugs.^16^ This last factor is considered to be a major determinant of risk associated with drug-induced LQTS when torsadogenic drugs are co-administered with other drugs in patients with polypharmacy. On one hand, it is often stated that combined administration of I_Kr_ blocking drugs (expected synergistic pharmacodynamic effects) could lead to increased QT prolongation. Although this appears to be the case, our studies have demonstrated that concomitant block of I_Kr_ was not necessarily associated with synergistic or potentiation of drug effects.^46^ However, our team has shown that a combined block of I_Kr_ and I_Ks_ was associated with potentiation of drug effects on cardiac repolarization.^47^ On the other hand, the role of competitive inhibition (pharmacokinetic interactions) due to inhibition of the metabolism of the torsadogenic drug, leading to its increased systemic exposure, is recognized but often overlooked.^48^ For the Long QT-JT index, a value ≤15 is associated with an increased risk of QT prolongation and induction of TdP by the drug (“High Risk”), while a value >15 and ≤100 is associated with an increased risk of QT prolongation (“Moderate Risk”); values >101 are qualified as “Low Risk”.

Arrhythmogenic effects of COVID-19 drugs could be expected, potentially contributing to disease outcome.^6^ This may be of importance for patients with an increased risk for cardiac arrhythmias, either secondary to acquired conditions or co-morbidities or consequent to inherited syndromes.^49^ Especially, patients with hepatic diseases, heart failure, renal failure and electrolyte disturbances are at increased risk of drug-induced LQTS. In such context, taking into consideration patients’ characteristics, various algorithms have been reported to identify patients that are more susceptible to experiencing drug induced QT prolongation.^50^

Experimental COVID-19 use with mandatory monitoring of the QT interval is defensible because the benefits of using some of the proposed drugs could outweigh the risks. Key considerations supporting their use are:

1. The duration of use for these medications in COVID-19 is shorter than their original indication (chronic *vs* five to ten days) thus, only a short-term monitoring would be required.
2. The overall potential population-benefits of those drugs if proven to be effective for COVID-19 compared to the number of patients at high risk for QT prolongation.

In addition to monitoring of the QT interval, samples should be collected and a biobank created to support research efforts in the present and future. There is an urgent need for long-term, placebo controlled, randomized clinical trials. It is recognized that polymorphisms in genes regulating the pharmacokinetic and pharmacodynamic pathways of these drugs may impact an individual’s drug response and consequently, the overall outcomes.

The risk-benefit assessment for the use of repurposed drugs for the treatment of COVID-19 remains unclear since benefits are currently awaited, not proven, but our search strategy indicates that several of these drugs show significant risk of toxicity, especially drug-induced LQTS, and adverse drug events.

## Data Availability

The authors confirm that the data supporting the findings of this study are available within the article and/or its supplementary materials.

https://www.trhc.spprdi.com/covid19/home

## STUDY HIGHLIGHTS

- Our study systematically explores current research and validated indices related to drug-induced LQTS, in addition to consider the various mechanisms of action for these drugs to induce LQTS.
- To our knowledge, this is the first comparison of risk for a prolonged QT interval and torsade de pointes in the use of hydroxychloroquine, chloroquine, remdesivir or lopinavir/ritonavir to 23 well-known torsadogenic drugs.
- We also present a comprehensive analysis of arrhythmogenic events reported for COVID-19 drugs and torsadogenic compounds from the FAERS database.
- Our conclusions are limited by the nature of data obtained from a literature search and extraction from databases, limited availability of basic pharmacokinetic and pharmacodynamic parameters and clinical data for some of the newer COVID-19 drugs.

## Author Contributions

VM, JT, and PD wrote the manuscript; VM, PD, SBR, MD, MA, BC, and JT designed the research; SBR, MD, MA, PD, and BC performed the research. MD, MA, and SBR analyzed the data.

## Acknowledgements

The authors want to thank Dana Filippoli and Dr. Calvin H. Knowlton for review and insightful comments on the content of the paper. The authors also recognize the contribution of Ernesto Lucio, Gerald Condon, Ravil Bikmetov, PhD, and Matt K Smith, PhD.

## Conflict of Interest

Jacques Turgeon, Veronique Michaud, Pamela Dow, Sweilem Al Rihani, Malavika Deodhar, and Meghan Arwood are employees of Tabula Rasa HealthCare. Brian Cicali is a former employee of Tabula Rasa HealthCare and is currently an independent contractor. All authors possess or have possessed shares of Tabula Rasa HealthCare.

## Funding

Funding for this research was made possible by Tabula Rasa HealthCare.

## Notes

### Competing Interest Statement

Veronique Michaud, Pamela Dow, Sweilem Al Rihani, Malavika Deodhar, Meghan Arwood, and Jacques Turgeon are all employees of Tabula Rasa HealthCare. Jacques Turgeon and Veronique Michaud are stock holders of Tabula Rasa HealthCare. Jacques Turgeon and Veronique Michaud have patents broadly related to the content of this paper. Veronique Michaud and Jacques Turgeon are faculty members at the Universite de Montreal. Brian Cicali is a previous employee of Tabula Rasa HealthCare, and is paid independently by TRHC for work outside of this paper.

### Funding Statement

This research was made possible by funding from Tabula Rasa HealthCare.

